# Trans-ancestry genetic study of type 2 diabetes highlights the power of diverse populations for discovery and translation

**DOI:** 10.1101/2020.09.22.20198937

**Authors:** Anubha Mahajan, Cassandra N Spracklen, Weihua Zhang, Maggie CY Ng, Lauren E Petty, Hidetoshi Kitajima, Grace Z Yu, Sina Rüeger, Leo Speidel, Young Jin Kim, Momoko Horikoshi, Josep M Mercader, Daniel Taliun, Sanghoon Moon, Soo-Heon Kwak, Neil R Robertson, Nigel W Rayner, Marie Loh, Bong-Jo Kim, Joshua Chiou, Irene Miguel-Escalada, Pietro della Briotta Parolo, Kuang Lin, Fiona Bragg, Michael H Preuss, Fumihiko Takeuchi, Jana Nano, Xiuqing Guo, Amel Lamri, Masahiro Nakatochi, Robert A Scott, Jung-Jin Lee, Alicia Huerta-Chagoya, Mariaelisa Graff, Jin-Fang Chai, Esteban J Parra, Jie Yao, Lawrence F Bielak, Yasuharu Tabara, Yang Hai, Valgerdur Steinthorsdottir, James P Cook, Mart Kals, Niels Grarup, Ellen M Schmidt, Ian Pan, Tamar Sofer, Matthias Wuttke, Chloe Sarnowski, Christian Gieger, Darryl Nousome, Stella Trompet, Jirong Long, Meng Sun, Lin Tong, Wei-Min Chen, Meraj Ahmad, Raymond Noordam, Victor JY Lim, Claudia HT Tam, Yoonjung Yoonie Joo, Chien-Hsiun Chen, Laura M Raffield, Cécile Lecoeur, Nisa M Maruthur, Bram Peter Prins, Aude Nicolas, Lisa R Yanek, Guanjie Chen, Richard A Jensen, Salman Tajuddin, Edmond Kabagambe, Ping An, Anny H Xiang, Hyeok Sun Choi, Brian E Cade, Jingyi Tan, Fernando Abaitua, Linda S Adair, Adebowale Adeyemo, Carlos A Aguilar-Salinas, Masato Akiyama, Sonia S Anand, Alain Bertoni, Zheng Bian, Jette Bork-Jensen, Ivan Brandslund, Jennifer A Brody, Chad M Brummett, Thomas A Buchanan, Mickaël Canouil, Juliana CN Chan, Li-Ching Chang, Miao-Li Chee, Ji Chen, Shyh-Huei Chen, Yuan-Tsong Chen, Zhengming Chen, Lee-Ming Chuang, Mary Cushman, Swapan K Das, H. Janaka de Silva, George Dedoussis, Latchezar Dimitrov, Ayo P Doumatey, Shufa Du, Qing Duan, Kai-Uwe Eckardt, Leslie S Emery, Daniel S Evans, Michele K Evans, Krista Fischer, James S Floyd, Ian Ford, Myriam Fornage, Oscar H Franco, Timothy M Frayling, Barry I Freedman, Christian Fuchsberger, Pauline Genter, Hertzel C Gerstein, Vilmantas Giedraitis, Clicerio González-Villalpando, Maria Elena González-Villalpando, Mark O Goodarzi, Penny Gordon-Larsen, David Gorkin, Myron Gross, Yu Guo, Sophie Hackinger, Sohee Han, Andrew T Hattersley, Christian Herder, Annie-Green Howard, Willa Hsueh, Mengna Huang, Wei Huang, Yi-Jen Hung, Mi Yeong Hwang, Chii-Min Hwu, Sahoko Ichihara, Mohammad Arfan Ikram, Martin Ingelsson, Md. Tariqul Islam, Masato Isono, Hye-Mi Jang, Farzana Jasmine, Guozhi Jiang, Jost B Jonas, Marit E Jørgensen, Torben Jørgensen, Yoichiro Kamatani, Fouad R Kandeel, Anuradhani Kasturiratne, Tomohiro Katsuya, Varinderpal Kaur, Takahisa Kawaguchi, Jacob M Keaton, Abel N Kho, Chiea-Chuen Khor, Muhammad G Kibriya, Duk-Hwan Kim, Katsuhiko Kohara, Jennifer Kriebel, Florian Kronenberg, Johanna Kuusisto, Kristi Läll, Leslie A Lange, Myung-Shik Lee, Nanette R Lee, Aaron Leong, Liming Li, Yun Li, Ruifang Li-Gao, Symen Ligthart, Cecilia M Lindgren, Allan Linneberg, Ching-Ti Liu, Jianjun Liu, Adam E Locke, Tin Louie, Jian’an Luan, Andrea O Luk, Xi Luo, Jun Lv, Valeriya Lyssenko, Vasiliki Mamakou, K Radha Mani, Thomas Meitinger, Andres Metspalu, Andrew D Morris, Girish N. Nadkarni, Jerry L Nadler, Michael A Nalls, Uma Nayak, Ioanna Ntalla, Yukinori Okada, Lorena Orozco, Sanjay R Patel, Mark A Pereira, Annette Peters, Fraser J Pirie, Bianca Porneala, Gauri Prasad, Sebastian Preissl, Laura J Rasmussen-Torvik, Alexander P Reiner, Michael Roden, Rebecca Rohde, Katheryn Roll, Charumathi Sabanayagam, Maike Sander, Kevin Sandow, Naveed Sattar, Sebastian Schönherr, Claudia Schurmann, Mohammad Shahriar, Jinxiu Shi, Dong Mun Shin, Daniel Shriner, Jennifer A Smith, Wing Yee So, Alena Stančáková, Adrienne M Stilp, Konstantin Strauch, Ken Suzuki, Atsushi Takahashi, Kent D Taylor, Barbara Thorand, Gudmar Thorleifsson, Unnur Thorsteinsdottir, Brian Tomlinson, Jason M Torres, Fuu-Jen Tsai, Jaakko Tuomilehto, Teresa Tusie-Luna, Miriam S Udler, Adan Valladares-Salgado, Rob M van Dam, Jan B van Klinken, Rohit Varma, Marijana Vujkovic, Niels Wacher-Rodarte, Ellie Wheeler, Eric A Whitsel, Ananda R Wickremasinghe, Konstantin Willems van Dijk, Daniel R Witte, Chittaranjan S Yajnik, Ken Yamamoto, Toshimasa Yamauchi, Loïc Yengo, Kyungheon Yoon, Canqing Yu, Jian-Min Yuan, Salim Yusuf, Liang Zhang, Wei Zheng, FinnGen, Leslie J Raffel, Michiya Igase, Eli Ipp, Susan Redline, Yoon Shin Cho, Lars Lind, Michael A Province, Craig L Hanis, Patricia A Peyser, Erik Ingelsson, Alan B Zonderman, Bruce M Psaty, Ya-Xing Wang, Charles N Rotimi, Diane M Becker, Fumihiko Matsuda, Yongmei Liu, Eleftheria Zeggini, Mitsuhiro Yokota, Stephen S Rich, Charles Kooperberg, James S Pankow, James C Engert, Yii-Der Ida Chen, Philippe Froguel, James G Wilson, Wayne HH Sheu, Sharon LR Kardia, Jer-Yuarn Wu, M Geoffrey Hayes, Ronald CW Ma, Tien-Yin Wong, Leif Groop, Dennis O Mook-Kanamori, Giriraj R Chandak, Francis S Collins, Dwaipayan Bharadwaj, Guillaume Paré, Michèle M Sale, Habibul Ahsan, Ayesha A Motala, Xiao-Ou Shu, Kyong-Soo Park, J Wouter Jukema, Miguel Cruz, Roberta McKean-Cowdin, Harald Grallert, Ching-Yu Cheng, Erwin P Bottinger, Abbas Dehghan, E-Shyong Tai, Josee Dupuis, Norihiro Kato, Markku Laakso, Anna Köttgen, Woon-Puay Koh, Colin NA Palmer, Simin Liu, Goncalo Abecasis, Jaspal S Kooner, Ruth JF Loos, Kari E North, Christopher A Haiman, Jose C Florez, Danish Saleheen, Torben Hansen, Oluf Pedersen, Reedik Mägi, Claudia Langenberg, Nicholas J Wareham, Shiro Maeda, Takashi Kadowaki, Juyoung Lee, Iona Y Millwood, Robin G Walters, Kari Stefansson, Simon R Myers, Jorge Ferrer, Kyle J Gaulton, James B Meigs, Karen L Mohlke, Anna L Gloyn, Donald W Bowden, Jennifer E Below, John C Chambers, Xueling Sim, Michael Boehnke, Jerome I Rotter, Mark I McCarthy, Andrew P Morris, on behalf of the DIAMANTE Consortium

## Abstract

We assembled an ancestrally diverse collection of genome-wide association studies of type 2 diabetes (T2D) in 180,834 cases and 1,159,055 controls (48.9% non-European descent). We identified 277 loci at genome-wide significance (*p*<5×10^-8^), including 237 attaining a more stringent trans-ancestry threshold (*p*<5×10^-9^), which were delineated to 338 distinct association signals. Trans-ancestry meta-regression offered substantial enhancements to fine-mapping, with 58.6% of associations more precisely localised due to population diversity, and 54.4% of signals resolved to a single variant with >50% posterior probability. This improved fine-mapping enabled systematic assessment of candidate causal genes and molecular mechanisms through which T2D associations are mediated, laying foundations for functional investigations. Trans-ancestry genetic risk scores enhanced transferability across diverse populations, providing a step towards more effective clinical translation to improve global health.

## MAIN TEXT

Type 2 diabetes (T2D) is increasing in prevalence worldwide and contributes substantially to the global burden of mortality and disability. Genome-wide association studies (GWAS) have been extremely successful in identifying T2D susceptibility loci but have mostly focused on populations of European ancestry (*1-4*). To address this population bias, more recent large-scale T2D GWAS have expanded into East Asian ancestry populations (*5,6*) and multi-ancestry biobanks (*7,8*). The DIAMANTE (DIAbetes Meta-ANalysis of Trans-Ethnic association studies) Consortium was established to assemble T2D GWAS across diverse populations and develop methodological blueprints for intra- and inter-ancestry meta-analyses relevant to any complex trait. Analyses of the European and East Asian ancestry-specific components of DIAMANTE have previously been reported (*4,6*). Here, we describe the results of our trans-ancestry meta-analysis of T2D GWAS, which partially overlaps with these ancestry-specific efforts, focussing on genetic variation shared across population groups at loci robustly associated with the disease. With these data, we demonstrate the value of analyses conducted in diverse populations to understand how T2D-associated variants impact downstream molecular and biological processes underlying the disease, and to advance clinical translation for all, irrespective of ancestry.

### Robust discovery of trans-ancestry T2D associations

We accumulated summary statistics from 122 GWAS with a total of 180,834 T2D cases and 1,159,055 controls (effective sample size 492,191) from five ancestry groups (**Figure S1, Tables S1 and S2**): European (51.1%); East Asian (28.4%); South Asian (8.3%); African (6.6%); and Hispanic/Latino (5.6%). Here, we use the term “ancestry group” to refer to populations with similar genetic ancestry and/or admixture profiles. We imputed each GWAS to reference panels from the 1000 Genomes Project (*9,10*), Haplotype Reference Consortium (*11*), or population-specific whole-genome sequence data (**Supplementary Materials and Methods, Table S3**). We considered 19,829,461 bi-allelic autosomal single nucleotide variants (SNVs) that overlapped the reference panels and had minor allele frequency (MAF) >0.5% in at least one of the five ancestry groups (**Supplementary Materials and Methods**).

The most powerful methods for discovery of novel loci through trans-ancestry meta-analysis allow for potential allelic effect heterogeneity between ancestry groups that cannot be accommodated in a fixed-effects model (*12*). In contrast, random-effects meta-analysis does not assume any structure to heterogeneity across GWAS. To address these limitations, our primary analysis used MR-MEGA (*13*), a meta-regression approach that models allelic effect heterogeneity correlated with ancestry by including axes of genetic variation as covariates to represent diversity between GWAS (**Supplementary Materials and Methods**). We considered three axes of genetic variation that separated GWAS from the five ancestry groups, and which also revealed finer-scale genetic differences between GWAS of similar ancestry (**Figure S2**). In particular, the second axis highlights the extent of African and European admixture amongst African American GWAS, whilst the third axis differentiates Hispanic/Latino studies with variable proportions of indigenous American and African ancestry.

We identified 277 loci associated with T2D at the traditional genome-wide significance threshold of *p*<5×10^-8^ in the trans-ancestry meta-regression (**Figure S3, Table S4**). By accounting for ancestry-correlated allelic effect heterogeneity in the trans-ancestry meta-regression, we observed lower genomic control inflation (λ_GC_=1.05) than when using either fixed- or random-effects meta-analysis (λ_GC_=1.25 under both models). Consequently, after genomic control correction, we observed stronger signals of association at lead SNVs at most loci in the meta-regression than in the fixed- or random-effects meta-analyses (**Figure S4**). Eleven of the 277 loci have not been reported in recently published T2D GWAS meta-analyses (*4,6,8*), which together contributed 78.6% of the total effective sample size of this trans-ancestry meta-regression (**Figure S3**). **Supplementary Note 1** summarises the loci reported across investigations incorporating DIAMANTE GWAS.

To gain insight into the power offered by aggregating GWAS from diverse populations, we sought to compare the number of loci identified in trans-ancestry meta-regression and ancestry-specific meta-analyses. There are minor differences in the GWAS contributing to the trans-ancestry meta-regression and the previously reported European and East Asian ancestry-specific components of DIAMANTE (*4,6*). To make a direct comparison of locus discovery, we therefore undertook ancestry-specific fixed-effects meta-analyses of only those GWAS contributing to the trans-ancestry meta-regression (**Supplementary Materials and Methods**). Of the 100 and 193 loci attaining genome-wide significance in the East Asian and European ancestry-specific meta-analyses, respectively, lead SNVs at 94 (94.0%) and 164 (85.0%) demonstrated stronger evidence for association in the trans-ancestry meta-regression (**Figure S5**). In contrast, eleven (5.7%) of the 193 loci identified in the European ancestry-specific meta-analysis did not attain genome-wide significance in the trans-ancestry meta-regression (**Table S5**). The lead SNVs at these loci demonstrated no significant differences in allelic effect sizes across European ancestry GWAS (Cochran’s *Q p*<0.0045, Bonferroni correction), which would suggest that these signals are not false positive associations. None of these SNVs demonstrated significant evidence of T2D association in a meta-analysis of non-European ancestry GWAS, indicating that they could represent European ancestry-specific loci. Such signals could arise when the lead SNV is in strong linkage disequilibrium (LD) with an ancestry-specific causal variant that has not been interrogated in the trans-ancestry meta-regression. Taken together, these results demonstrate the power of diverse populations for locus discovery and replication, but also emphasize the importance of single-ancestry GWAS for optimal identification of ancestry-specific associations.

The traditional genome-wide significance threshold of *p*<5×10^-8^ was derived from a Bonferroni correction for the equivalent of approximately one million independent, common SNVs in European ancestry populations. However, in a trans-ancestry meta-analysis, the correlation in association summary statistics between SNVs will be weaker because of the different patterns of LD across diverse populations. We therefore derived a more stringent trans-ancestry genome-wide significance threshold of *p*<5×10^-9^ by estimating the effective number of independent SNVs across ancestries using haplotypes from the 1000 Genomes Project reference panel (*10*) (**Supplementary Materials and Methods**). Of the 277 loci reported in this trans-ancestry meta-regression, 237 attained the more stringent significance threshold, including the novel associations mapping to/near *PXK, PPP3CA, MYO3A* and *FOLH1*. We focussed only on these 237 loci for the downstream analyses presented here.

Through approximate conditional analyses, conducted using ancestry-matched LD reference panels for each GWAS, we partitioned associations at the 237 T2D loci into 338 distinct signals that were each represented by an index SNV at trans-ancestry genome-wide significance (**Supplementary Materials and Methods, Tables S6 and S7**). We observed multiple distinct association signals at 52 (21.9%) loci, of which 50 were represented by between two and five index SNVs. The most complex genetic architecture was observed across a 1Mb region flanking the lead SNV at the *TCF7L2* locus, where the T2D association was delineated to 16 distinct signals, and a 1.7Mb imprinted region encompassing the previously reported loci *INS-IGF2* and *KCNQ1*, which was decomposed into 14 distinct signals (**Figures S6 and S7**).

### Extent and source of trans-ancestry allelic-effect heterogeneity

Of the 338 distinct T2D signals, 317 (93.8%) attained at least nominal evidence of association (*p*<0.05) in two or more ancestry groups (**Table S8**). The strength of association (*p*-value) in each ancestry depends on the effective sample size, and the frequency and effect size (odds-ratio, OR) of the risk allele at the index SNV (**Figure S8**). Trans-ancestry heterogeneity in allelic effects at an association signal can occur for several reasons, including differences in LD with the causal variant and/or interaction with lifestyle factors that vary across diverse populations. An advantage of the MR-MEGA meta-regression model is that heterogeneity can be partitioned into two components. The first captures heterogeneity that is correlated with ancestry (i.e. can be explained by the three axes of genetic variation). The second reflects residual heterogeneity due to differences in study design (for example variable phenotype definition, case-control ascertainment, or covariate adjustments between GWAS). We observed 27 (8.0%) distinct T2D associations with nominal evidence (*p*_*HET*_<0.05) of residual heterogeneity compared to that expected by chance (binomial test *p*=0.0037). In contrast, there was nominal evidence of ancestry-correlated heterogeneity at 136 (40.2%) T2D association signals (binomial test *p*<2.2×10^-16^), suggesting that differences in allelic effect sizes between GWAS are more likely due to factors related to ancestry than to study design.

We next sought to investigate whether these ancestry-correlated heterogeneous association signals could be explained by an interaction with obesity, given that the distribution of body mass index (BMI) is shifted to the left in individuals of East Asian ancestry. To do this, we considered the 136 association signals with nominal evidence of ancestry-correlated heterogeneity in allelic effects, and repeated the meta-regression analysis at each index SNV by including the mean BMI of individuals in each study as a covariate in addition to the three axes of genetic variation (**Supplementary Materials and Methods, Table S9**). The strongest evidence for heterogeneity in allelic effects that was correlated with BMI (after accounting for ancestry) was observed for the T2D association signals at the *CDKAL1* locus (rs9348441, *p*_HET_=3.0×10^-6^). At this signal, the effect of the risk allele on T2D was greatest in East Asian ancestry populations, and there was a negative correlation between BMI and log-OR across GWAS (**Figure S9**). This relationship is consistent with the notion of “favourable adiposity”, where BMI increasing alleles are associated with lower insulin levels and higher subcutaneous-to-visceral adipose tissue ratio, which may protect against T2D via higher adipose storage capacity (*14*). A potential protective interaction with obesity at this locus is supported by: (i) evidence of a stronger association at the index SNV after adjustment for BMI in the European and East Asian ancestry-specific components of DIAMANTE (*4,6*); and (ii) the T2D-risk allele is significantly associated with decreased BMI in European and East Asian ancestry GWAS meta-analyses of obesity in the general population (*15,16*).

### Population diversity improves fine-mapping resolution

The value of exploiting differences in LD structure between ancestry groups to enhance the localisation of causal variants has been demonstrated in previous T2D GWAS meta-analyses (*17,18*). We sought to quantify the improvement in fine-mapping resolution offered by increased sample size and population diversity in the trans-ancestry meta-regression over the European ancestry-specific meta-analysis (since the latter represents the largest ancestry-specific component of DIAMANTE). For each of the 338 distinct signals, we derived trans-ancestry and European ancestry-specific credible sets of variants that account for 99% of the posterior probability (*π*) of driving the T2D association under a uniform prior model of causality (**Supplementary Materials and Methods**). Trans-ancestry meta-regression substantially reduced the median 99% credible set size from 35 variants (spanning 112kb) to 10 variants (spanning 26kb), and increased the median posterior probability ascribed to the index SNV from 24.3% to 42.0%. The 99% credible sets for 266 (78.7%) distinct T2D associations were smaller in the trans-ancestry meta-regression than in the European ancestry-specific meta-analysis, whilst a further 26 (7.7%) signals were resolved to a single SNV in both (**Figure 1, Table S10**).

**Figure 1.**
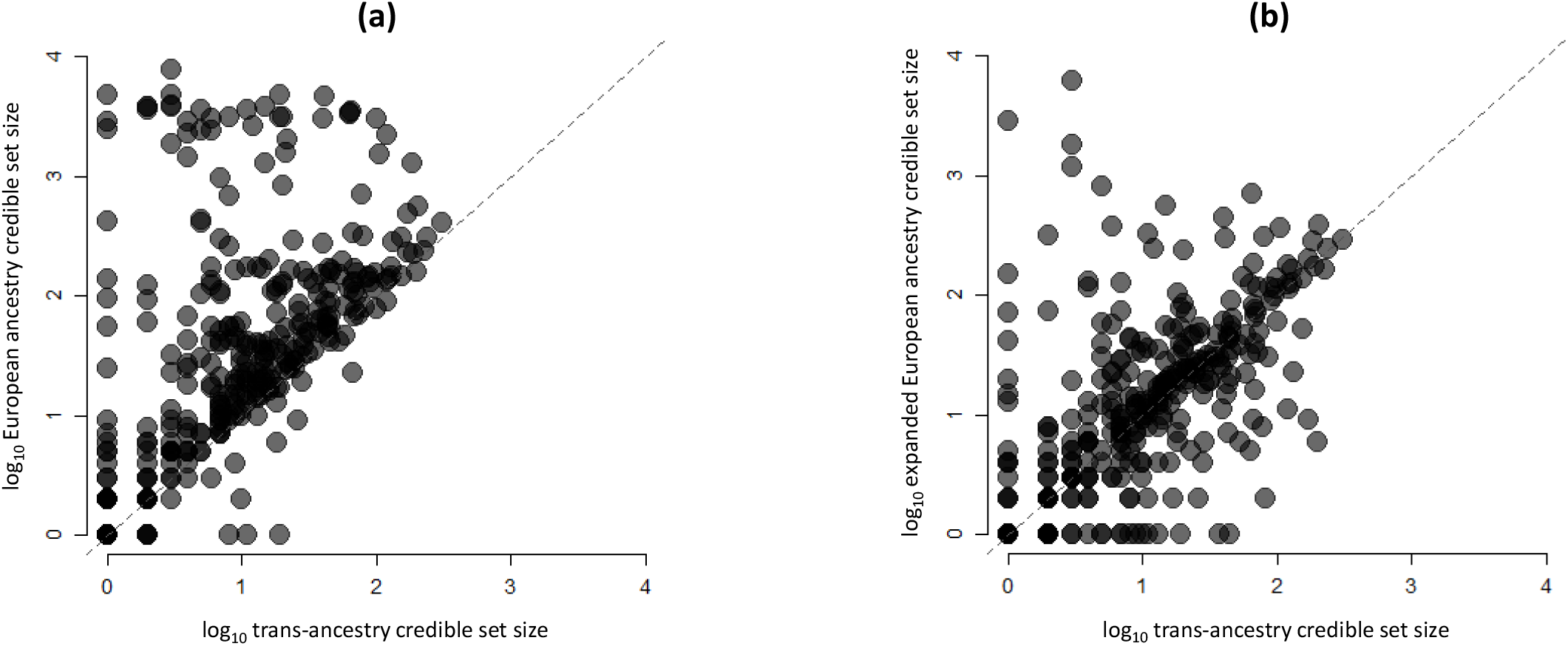
Comparison of number of SNVs in 99% credible set for distinct association signals for T2D obtained from European ancestry-specific meta-analysis and trans-ancestry meta-regression. Each point corresponds to a distinct association signal, plotted according to the log_10_ credible set size in the trans-ancestry meta-regression on the x-axis and the log_10_ credible set size in the European ancestry meta-analysis on the y-axis. Trans-ancestry credible sets were derived from meta-regression of 180,834 cases and 1,159,055 controls. European ancestry-specific credible sets were derived from meta-analysis of 80,154 cases and 853,616 controls. (a) We excluded 26 signals that were resolved to a single variant in both trans-ancestry meta-regression and European ancestry-specific meta-analysis. The 266 (85.3%) signals above the dashed y=x line were more precisely fine-mapped in the trans-ancestry meta-regression. (b) We artificially boosted the effective sample size of the European ancestry-specific meta-analysis to that of the trans-ancestry meta-regression. We considered the 266 signals that were more precisely fine-mapped in the trans-ancestry meta-regression. The 156 (58.6%) signals above the dashed y=x line were more precisely fine-mapped because of population diversity.

Compared to the European ancestry-specific meta-analysis, some of the most dramatic improvements in fine-mapping resolution after trans-ancestry meta-regression included signals where the index SNV was of lower frequency and/or of smaller effect in European ancestry populations. For these signals, including those at *GCC1-PAX4-LEP, SGCG, RGMA, DSTYK-MDM4* and *MYO3A*, the evidence for association was weak in the European ancestry-specific meta-analysis, resulting in large credible sets compared to other ancestry groups. However, we also observed examples of T2D signals with strong associations across all five ancestry groups for which the credible sets were smaller in the trans-ancestry meta-regression, with the most noticeable improvements in fine-mapping resolution observed at *TMEM154, HMGA2, GRP-MC4R, IGF2BP2, SPRY2* and *FTO* (**Table S10**). At *FTO*, the 18 variants in the European ancestry-specific 99% credible set were in strong LD with the index SNV (rs55872725) in European ancestry populations (*r*^2^>0.8). However, the 99% credible set after trans-ancestry meta-regression included just six of these variants that were in strong LD with the index SNV in all five ancestry groups (**Figure S10**).

We next attempted to understand the relative contributions of population diversity and sample size to these improvements in fine-mapping resolution. We artificially boosted the effective sample size of the European ancestry-specific meta-analysis to that of the trans-ancestry meta-regression by recalibrating standard errors of allelic effect estimates and the genomic control inflation factor (**Supplementary Materials and Methods**). We considered the 266 distinct T2D associations that were more precisely localised after trans-ancestry meta-regression. Of these, the 99% credible sets at 156 (58.6%) signals were better resolved in the trans-ancestry meta-regression (**Figure 1, Table S10**). At these signals, the addition of non-European ancestry GWAS to the trans-ancestry meta-regression offered greater fine-mapping resolution than equivalent-sized European descent studies. These results highlight the value of diverse populations for causal variant localisation, emphasizing the importance of differences in LD structure and allele frequency distribution between ancestry groups.

### Annotation-informed trans-ancestry fine-mapping deconvolutes distinct T2D associations to single variant resolution

Previous T2D GWAS have demonstrated improved localisation of causal variants through integration of fine-mapping data with genomic annotation (*4,19*). By mapping SNVs to three categories of functional and regulatory annotation, with an emphasis on diabetes-relevant tissues (*20*), we observed significant joint enrichment (*p*<0.00023, Bonferroni correction for 220 annotations) for T2D associations mapping to protein coding exons, binding sites for NKX2.2, FOXA2, EZH2 and PDX1, and four chromatin states in pancreatic islets that mark active enhancers, active promoters and transcribed regions (**Supplementary Materials and Methods, Table S11, Figure S11**). We used the enriched annotations to derive a prior model for causality, and redefined 99% credible sets of variants for each distinct signal (**Supplementary Materials and Methods, Table S12**). Annotation-informed fine-mapping reduced the size of the 99% credible set, compared to the uniform prior, at 144 (42.6%) distinct association signals (**Figure S12**), decreasing the median from 10 variants (spanning 26kb) to 8 variants (spanning 23kb). For 98 (29.0%) signals, a single SNV accounted for >90% of the posterior probability of the T2D association (**Table S13**). At a further 86 (25.4%) signals, >50% of the posterior probability could be attributed to a single SNV.

### Missense variants driving T2D associations implicate candidate causal genes

After annotation-informed trans-ancestry fine-mapping, 19 of the 184 SNVs accounting for >50% of the posterior probability of the T2D association were missense variants (**Table S14**). These included two that implicate novel candidate causal genes for the disease: *MYO5C* p.Glu1075Lys (rs3825801, *p*=3.8×10^-11^, *π*=69.2%) at the *MYO5C* locus; and *ACVR1C* p.Ile482Val (rs7594480, *p*=4.0×10^-12^, *π*=95.2%) at the *CYTIP* locus. *ACVR1C* encodes ALK7, a transforming growth factor-β receptor, overexpression of which induces growth inhibition and apoptosis of pancreatic β-cells (*21*). Three *ACVR1C* missense variants have been previously associated with body fat distribution (*22*).

The trans-ancestry meta-regression also highlighted two examples of previously reported T2D coding variant associations that were better resolved by fine-mapping across diverse populations (**Figure S13**). A set of five coding variants in *SLC16A11* has been associated with T2D in populations of Hispanic/Latino ancestry (*23,24*), but causality could not be ascribed because of strong LD between them. However, after trans-ancestry fine-mapping, *SLC16A11* p.Val113Ile (rs117767867, *p*=6.5×10^-24^, *π*=59.8%) emerged as the variant most likely driving this association signal (the other four coding variants together account for just 14.0% of the posterior probability). Similarly, strong LD at the *KCNJ11-ABCC8* locus has frustrated efforts in European ancestry studies to distinguish the impact on T2D of three missense variants: *KCNJ11* p.Val250Ile (rs5215), *KCNJ11* p.Lys23Glu (rs5219) and *ABCC8* p.Ala1369Ser (rs757110). *ABCC8* and *KCNJ11* code for the two elements of the hetero-octameric beta-cell K_ATP_ channel and both represent strong biological candidates. Whilst trans-ancestry fine-mapping cannot equivocally distinguish between *KCNJ11* p.Val250Ile (*p*=1.3×10^-54^, *π*=67.1%) and *KCNJ11* p.Lys23Glu (*p*=2.6×10^-54^, *π*=32.5%), the causal contribution of *ABCC8* p.Ala1369Ser (*p*=1.2×10^-51^, *π*=0.1%) to this association signal can be discounted.

Trans-ancestry fine-mapping also provided a more detailed view of the role of missense variants in driving three distinct T2D association signals at the *ZFAND3-KCNK16-GLP1R* locus (**Figure S14**). Previous East Asian ancestry GWAS and exome-array meta-analyses (*5,25*) reported T2D association with *GLP1R* p.Arg131Gln (rs3765467). Whilst this variant is included in the 99% credible set of the signal indexed by rs742762, a non-coding SNV, in the trans-ancestry meta-regression, it has a relatively low posterior probability of association (*π*=2.0%, compared with *π*=75.0% for the index SNV). However, we identified a different *GLP1R* missense variant, p.Pro7Leu (rs10305420, *p*=1.1×10^-9^, *π*=94.1%), not in LD with p.Arg131Gln, which seems likely to be causal for the second association signal at the locus. At the third signal, 61.4% of the posterior probability of association could be attributed to three different missense variants: p.Ser21Gly (rs10947804, *π*=39.2%) in *KCNK17*; and p.Pro254His (rs11756091, *π*=14.8%) and p.Ala277Glu (rs1535500, *π*=13.7%) in *KCNK16*. Both genes encode members of the TWIK-related alkaline pH-activated K2P family, TALK-1 and TALK-2, and are expressed in islets with high specificity. The missense variants are in strong LD with each other across ancestry groups, and the T2D-risk haplotype is associated with increased *KCNK17* expression in pancreatic islets (*20*). Taken together, these results highlight the complexity of coding variant associations with T2D at this locus, demonstrating a causal role for missense variants encoding *GLP1R, KCNK16* and *KCNK17* on disease risk.

### Multi-omics data integration highlights novel candidate effector genes and causal mechanisms for non-coding T2D association signals

We next sought to take advantage of the improved fine-mapping resolution offered by the trans-ancestry meta-regression to extend insight into candidate effector genes, tissue specificity and mechanisms through which regulatory variants at non-coding T2D association signals impact disease risk. Here, we describe how data from this study support further characterisation of the regulatory landscape through which T2D association signals are mediated, highlighting examples of two loci, *BCAR1* and *PROX1*, where multi-omics integration builds on our trans-ancestry fine-mapping to provide novel insights into the pathogenesis of the disease.

We first integrated annotation-informed fine-mapping data with molecular quantitative trait loci (QTL), *in cis*, for: (i) circulating plasma proteins (pQTL) (*26*); and (ii) gene expression (eQTL) in diverse tissues, including pancreatic islets, subcutaneous and visceral adipose, liver, skeletal muscle, and hypothalamus (*27,28*) (**Supplementary Materials and Methods**). Recognising that molecular QTL may also be driven by multiple causal variants, we dissected distinct plasma pQTL for each protein and distinct eQTL for each gene in each tissue via approximate conditional analyses. Bayesian colocalization (*29*) of each pair of distinct T2D associations and molecular QTL identified 97 candidate effector genes at 72 signals (posterior probability *π*_COLOC_ >80%): 7 *cis*-pQTL at 7 signals; and 126 *cis*-eQTL at 66 signals (**Tables S15 and S16**). These signals reinforced evidence supporting several genes that have been previously implicated in T2D through extensive functional studies, including *ADCY5, STARD10, IRS1, KLF14, SIX3* and *TCF7L2* (*30-34*). A single candidate effector gene was implicated at 49 T2D association signals, of which 10 colocalized with eQTL across multiple tissues: *CEP68, ITGB6, RBM6, PCGF3, JAZF1, ANK1, ABO, ARHGAP19, PLEKHA1* and *AP3S2*. In contrast, we observed that *cis*-eQTL at 44 signals were specific to one tissue (24 to pancreatic islets, 11 to subcutaneous adipose, five to skeletal muscle, two to visceral adipose, and one each to liver and hypothalamus), reiterating the importance of conducting colocalization analyses across multiple tissues.

We then intersected 99% credible set variants for distinct T2D association signals with genome-wide promoter-focussed chromatin conformation capture data (pcHi-C) from pancreatic islets, subcutaneous adipose and liver (not available in hypothalamus and visceral adipose) (*35-37*). Across the three tissues, we observed contacts between credible set variants and putative target gene promoters for 214 (63%) of the association signals (**Table S17**). The contacts at 119 of these signals were observed in only one tissue: 51 in islets, 45 in liver, and 23 in subcutaneous adipose. Some targets were expected based on their proximity to the index SNV for the T2D association (including *TCF7L2, PROX1, PTEN, DLEU1, GLIS3, CCND2, CMIP* and *BCL2*), but for 143 (67%) signals, we identified more distant candidate effector genes (including *AQP5* and *AQP6* at the *FAIM2* locus, *P2RX1* at the *ZZEF1* locus, *STX16* at the *GNAS* locus, and *ISL1* at the *ITGA1* locus). Several of these targets provided complementary support for candidate effector genes identified via colocalization with *cis*-eQTL in the same tissue: *PLEKHA1* in islets and subcutaneous adipose; *ST6GAL1, CARD9, DNLZ, CAMK1D, TCF7L2, TH, DLK1* and *AP3S2* in islets; *DCAF16, STEAP2* and *MAN2C1* in subcutaneous adipose; and *CEP68* and *SLC22A3* in liver.

At the *BCAR1* locus, trans-ancestry fine-mapping resolved the T2D association signal to a 99% credible set of nine variants. These variants overlap a chromatin accessible snATAC-seq peak in human pancreatic acinar cells *(38)* and an enhancer element in human pancreatic islets which interacts with an islet-accessible, active promoter upstream of the pancreatic exocrine enzyme chymotrypsin B2 gene *CTRB2 (36)*. The observations in bulk pancreatic islets are likely to have arisen due to exocrine (acinar cell) contamination since single cell data does not support its expression in endocrine cells (**Figure 2**). The T2D association signal also colocalized with a cis-pQTL for circulating plasma levels of chymotrypsin B1 (CTRB1, *π*_COLOC_=98.6%). Interestingly, by extending our colocalization analyses at this locus to *trans*-pQTL, we showed that variants driving the T2D association signal also act to regulate levels of three other pancreatic secretory enzymes produced by the acinar cells that are all involved in the digestion of ingested fats and proteins: carboxypeptidase B1 (CPB1, *π*_COLOC_=98.8%), pancreatic lipase related protein 1 (PLRP1, *π*_COLOC_=97.6%) and serine protease 2 (PRSS2, *π*_COLOC_=98.3%). These observations are consistent with an effect of the T2D-risk alleles on gene and protein expression in the exocrine pancreas, which subsequently influence pancreatic endocrine function. This is in line with a recent study reporting rare mutations in another protein produced by the exocrine pancreas, chymotrypsin-like elastase family member 2A, which were found to influence levels of digestive enzymes and glucagon, a glucose-raising hormone secreted from the alpha cells in the pancreatic islets *(39)*. Taken together, these complementary findings add to a growing body of evidence linking defects in the exocrine pancreas and T2D pathogenesis *(40,41)*.

**Figure 2.**
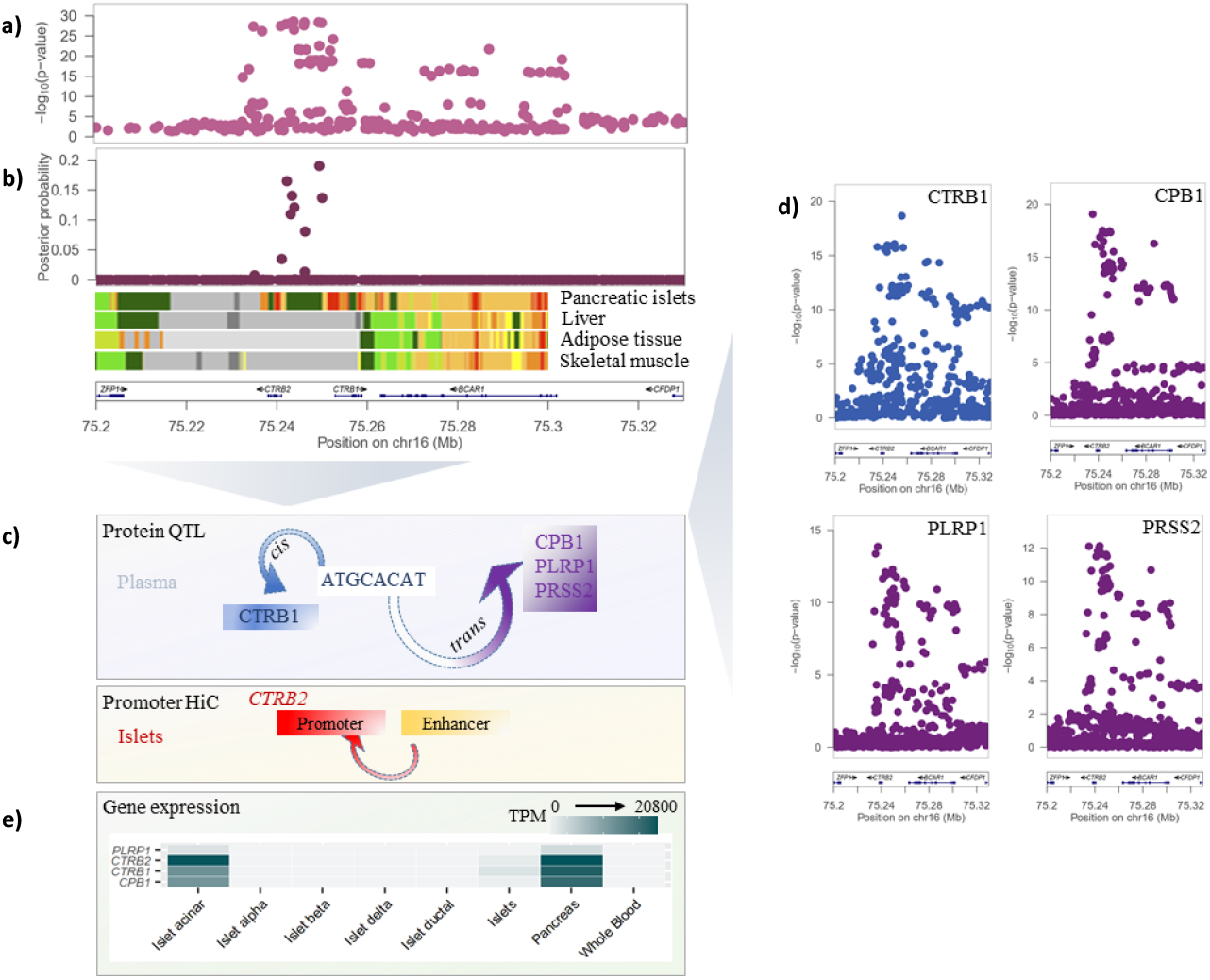
T2D association signal at the *BCAR1* locus colocalizes with multiple circulating plasma pQTLs. (a) Signal plot for T2D association from trans-ancestry meta-regression of 180,834 cases and 1,159,055 controls of diverse ancestry. Each point represents a SNV, plotted with their *p*-value (on a log_10_ scale) as a function of genomic position (NCBI build 37). Gene annotations are taken from the University of California Santa Cruz genome browser. Recombination rates are estimated from the Phase II HapMap. (b) Fine-mapping of T2D association signal from trans-ancestry meta-regression. Each point represents a SNV plotted with their posterior probability of driving T2D association as a function of genomic position (NCBI build 37). Chromatin states are presented for four diabetes-relevant tissues. (c) Schematic presentation of the single *cis*- and multiple *trans*-effects mediated by the *BCAR1* locus on plasma proteins and the islet chromatin loop between islet enhancer and promoter elements near *CTRB2*. (d) Signal plots for four circulating plasma proteins that colocalize with the T2D association in 3,301 European ancestry participants from the INTERVAL study. Each point represents a SNV, plotted with their *p*-value (on a log_10_ scale) as a function of genomic position (NCBI build 37). (e) Expression of genes (transcripts per million, TPM) encoding colocalized proteins in islets, pancreas and whole blood.

At the *PROX1* locus, trans-ancestry fine-mapping localised the two distinct association signals to just three variants (**Figure 3, Figure S15**). The index SNV at the first signal (rs340874, *p*=1.1×10^-18^, *π*>99.9%) overlaps the *PROX1* promoter in both human liver and pancreatic islets *(20,34)*. At the second signal, the two credible set variants are located in the same enhancer active in islets and liver (rs79687284, *p*=6.9×10^-19^, *π*=66.7%; rs17712208, *p*=1.4×10^-18^, *π*=33.3%). Recent studies have demonstrated that the T2D-risk allele at rs17712208 (but not rs79687284) results in significant repression of enhancer activity in mouse MIN6 *(38)* and human EndoC-βH1 beta cell models *(42)*. Furthermore, this enhancer interacts with the *PROX1* promoter in islets *(36)*, but not in liver *(37)*. Motivated by these observations, we sought to determine whether these distinct signals impact T2D risk (via *PROX1*) in a tissue-specific manner by assessing transcriptional activity of the credible set variants (rs340874, rs79687284 and rs17712208) in human HepG2 hepatocytes and EndoC-βH1 beta cell models using *in vitro* reporter assays (**Supplementary Materials and Methods, Figure 3**). At the first signal, we demonstrated significant differences in luciferase activity between alleles at rs340874 in both liver (*p*=0.0018) and islets (*p*=0.027). However, at the second signal, a significant difference in luciferase activity between alleles was observed only for rs17712208 in islets (*p*=0.00014). Taken together, these results suggest that likely causal variants at these distinct association signals exert their impact on T2D through the same effector gene, *PROX1*, but act in a tissue-specific manner.

**Figure 3.**
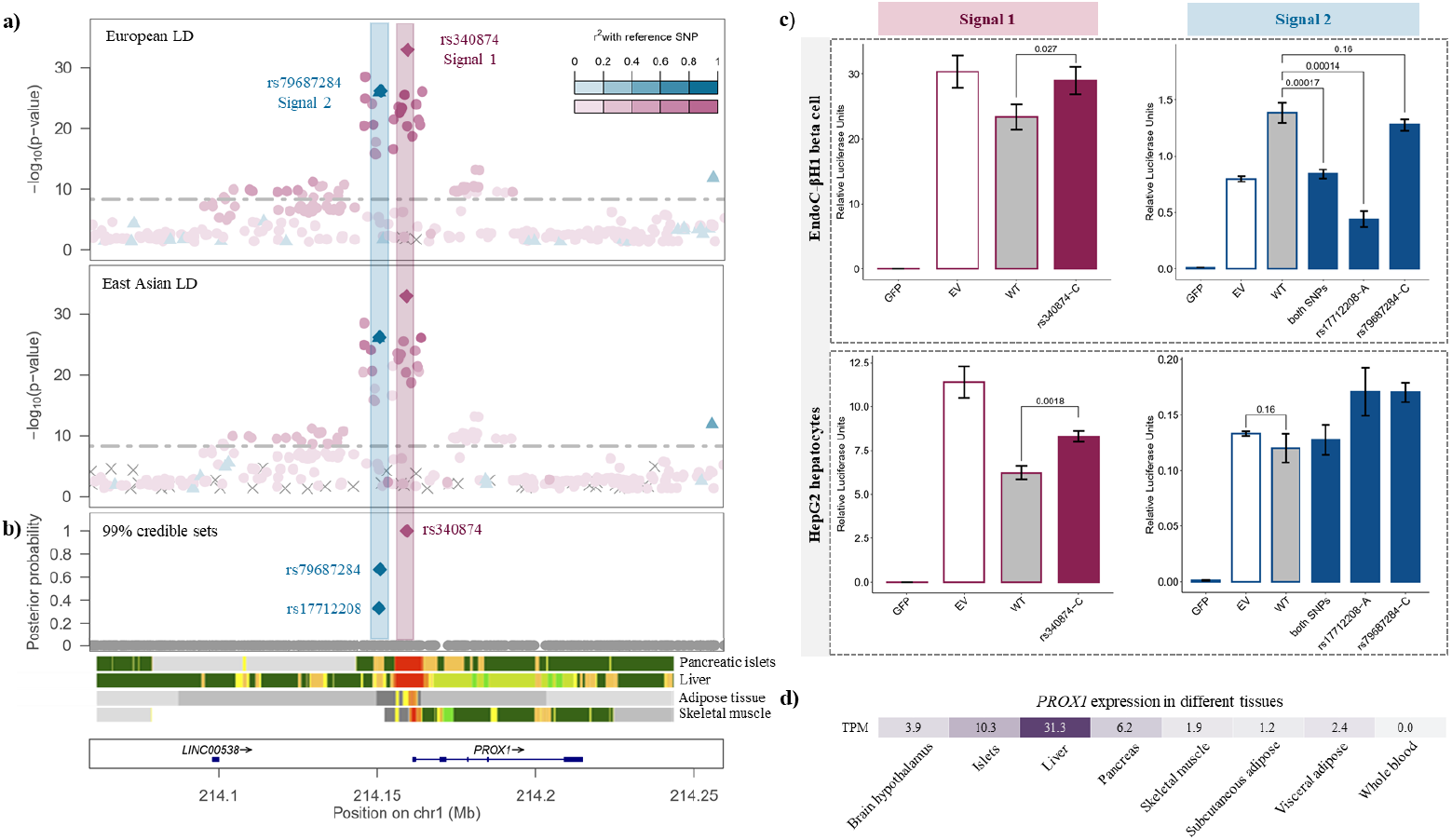
Defining causal molecular mechanisms at the *PROX1* locus. (a) Signal plot for two distinct T2D associations from trans-ancestry meta-regression of 180,834 cases and 1,159,055 controls of diverse ancestry. Each point represents a SNV, plotted with their *p*-value (on a -log_10_ scale) as a function of genomic position (NCBI build 37). Index SNVs are represented by the blue and purples diamonds. All other SNVs are coloured according the LD with the index SNVs in European and East Asian ancestry populations. Gene annotations are taken from the University of California Santa Cruz genome browser. (b) Fine-mapping of T2D association signals from trans-ancestry meta-regression. Each point represents a SNV plotted with their posterior probability of driving each distinct T2D association as a function of genomic position (NCBI build 37). The 99% credible sets for the two signals are highlighted by the purple and blue diamonds. Chromatin states are presented for four diabetes-relevant tissues. (c) Transcriptional activity of the 99 credible set variants at the two T2D association signals in human HepG2 hepatocytes and EndoC-βH1 beta cell models obtained from *in vitro* reporter assays. WT: wild-type (non-risk allele/haplotype). GFP: green fluorescent protein (negative control). EV: empty vector (baseline). (d) Expression of *PROX1* (transcripts per million, TPM) across a range of diabetes-relevant tissues.

### Transferability of T2D genetic risk scores across diverse populations

Genetic risk scores (GRS) derived from European ancestry GWAS have been demonstrated to have limited transferability into other population groups because of ancestry-correlated differences in the frequency and effect of risk alleles *(43)*. We took advantage of the population diversity in DIAMANTE to compare the prediction performance of trans-ancestry and ancestry-specific T2D GRS constructed using lead SNVs at loci attaining genome-wide significance. We hypothesized that the trans-ancestry GRS would perform best for two reasons. First, our analyses have highlighted that the trans-ancestry meta-regression enables discovery of loci associated with disease that are relevant across population groups. Second, we were able to take advantage of the meta-regression framework to estimate population-specific weights for each lead SNV in the GRS by allowing for ancestry-correlated heterogeneity in allelic effects (**Supplementary Materials and Methods**).

We began by selecting two studies per ancestry group as test GWAS into which the prediction performance of the GRS was assessed using trait variance explained (pseudo *R*^2^) and OR per risk score unit. We repeated the trans-ancestry meta-regression and ancestry-specific meta-analyses, after excluding the test GWAS, and defined lead SNVs at loci attaining genome-wide significance (*p*<5×10^-9^ for trans-ancestry GRS and *p*<5×10^-8^ for ancestry-specific GRS). For each ancestry-specific GRS, we used allelic effect estimates for each lead SNV as weights, irrespective of the population in which the test GWAS was undertaken. However, for the trans-ancestry GRS, we derived weights for each lead SNV that were specific to each test GWAS population. As expected, ancestry-specific GRS performed best into test GWAS from their respective ancestry group, but had relatively weak predictive power into other populations, particularly those of African descent (**Figure 4, Table S18**). However, the trans-ancestry GRS, weighted with population-specific allelic effect estimates from the meta-regression, showed improved predictive power over ancestry-specific GRS into all test GWAS.

**Figure 4.**
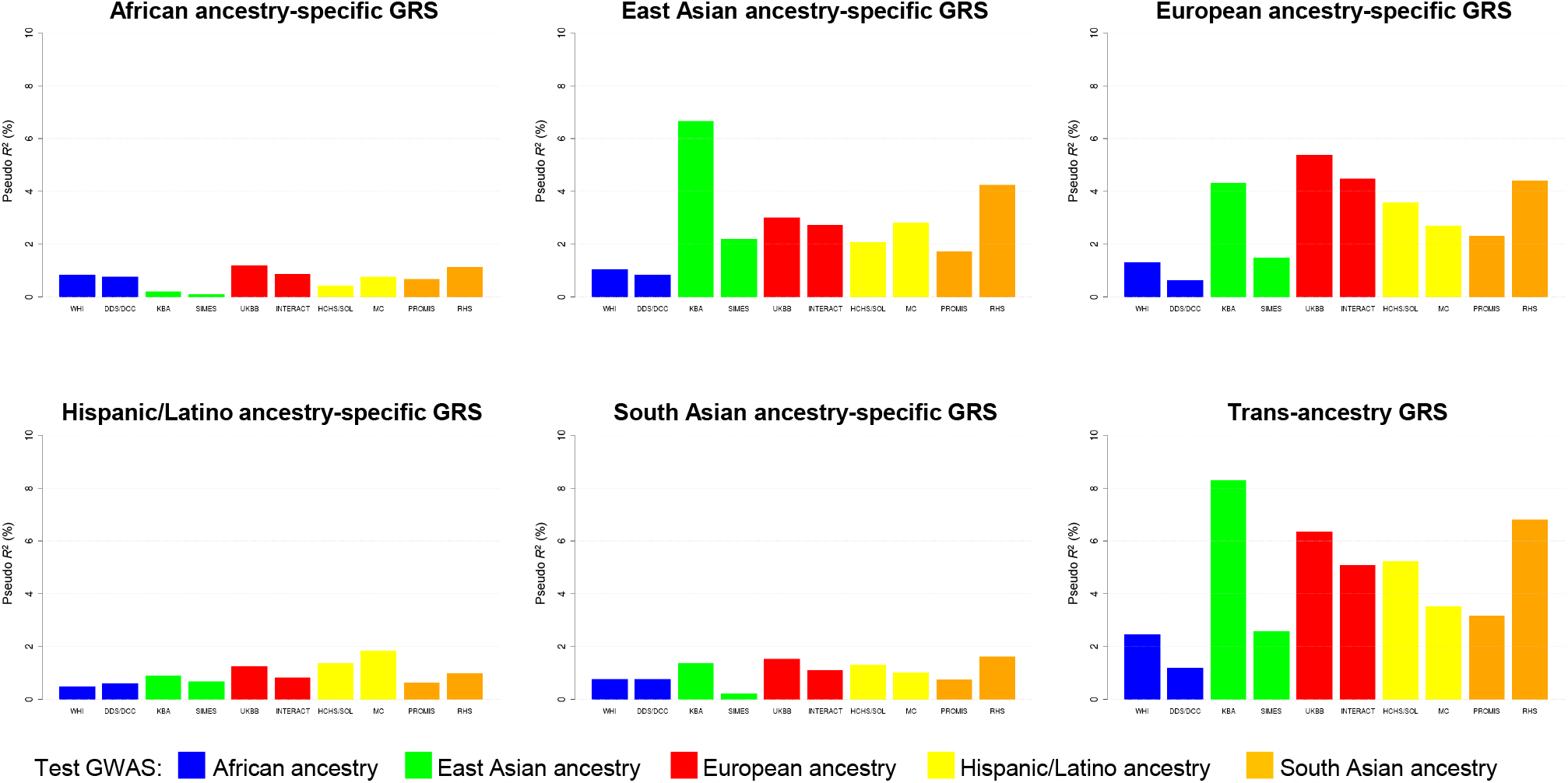
Transferability of trans-ancestry and ancestry-specific GRS into GWAS across diverse population groups. Each GRS was constructed using lead SNVs attaining genome-wide significance (*p*<5×10^-9^ for trans-ancestry GRS and *p*<5×10^-8^ for ancestry-specific GRS). For the trans-ancestry GRS, population-specific allelic effects on T2D were estimated from the meta-regression to generate different GRS weights for each test GWAS. For each ancestry-specific GRS, weights were generated from allelic effect estimates obtained from fixed-effects meta-analysis. The trait variance explained (pseudo *R*^2^) by each GRS was assessed in two test GWAS from each ancestry group. The trans-ancestry GRS out-performed ancestry-specific GRS into all test GWAS, reflecting the shared genetic contribution to T2D across diverse populations, despite differing allele frequencies and LD patterns.

The smallest increase in trait variance explained by the trans-ancestry GRS was observed into European ancestry test GWAS, in line with the substantial contribution of individuals of European descent to the trans-ancestry meta-regression. To further investigate the impact of sample size on prediction performance, we artificially boosted the effective sample size of the European ancestry-specific meta-analysis to that of the trans-ancestry meta-regression (after excluding test GWAS) using the same approach as in our fine-mapping analyses (**Supplementary Materials and Methods**). Expanding the sample size of the European ancestry-specific meta-analysis increased the number of loci attaining genome-wide significance: these corresponded to weaker association signals that were not discovered with the original sample size. Consequently, the trait variance explained by the European ancestry-specific GRS into test GWAS of European descent was increased with the expanded sample size. However, importantly, despite the increase in sample size, the trait variance explained did not exceed that attained by the trans-ancestry GRS (**Figure S16, Table S19**). In contrast, the trait variance explained by the European ancestry-specific GRS into test GWAS from non-European ancestry groups was lower with the expanded sample size than the original sample size. These results suggest that SNVs with weaker association signals in ancestry-specific meta-analyses are less likely to be transferable and thus reduce the predictive power of ancestry-specific GRS into other population groups.

Finally, we tested the predictive power of the trans-ancestry GRS based on all GWAS contributing to DIAMANTE into 18,111 T2D cases and 111,119 controls of Finnish descent from FinnGen (**Supplementary Materials and Methods**). Using association summary statistics from the meta-regression, we derived Finnish-specific allelic effect estimates to use as weights in the trans-ancestry GRS (**Figure S17, Table S20**). Individuals in the top decile of the GRS were at 5.34-fold increased risk of T2D compared to those in the bottom decile. The area under the receiver operating characteristic curve (AUC) of a predictive model including the Finnish-specific GRS, in addition to age, sex and BMI, was 83.5%. Each unit of the weighted GRS was associated with 1.24 years earlier age of T2D diagnosis (*p*=7.1×10^-57^), indicating that those with a higher genetic burden are more likely to be affected earlier in life.

### Positive selection of T2D risk alleles

Previous investigations *(44)* have concluded that historical positive selection has not had the major impact on T2D envisaged by the thrifty genotype hypothesis *(45)*. We sought to re-evaluate the evidence for positive selection of T2D risk alleles across our expanded collection of distinct trans-ancestry association signals. We fitted demographic histories to haplotypes for each population in the 1000 Genomes Project reference panel *(10)* using Relate *(46)*, and quantified the evidence for selection for each T2D index SNV by assessing the extent to which the mutation has more descendants than other lineages that were present when it arose (**Supplementary Materials and Methods**). This approach is well powered to detect positive selection acting on polygenic traits over a period of a few thousand to a few ten thousand years. We detected evidence of selection (*p*<0.05) in four of the five African ancestry populations (but not other ancestry groups) towards increased T2D risk (**Figure S18**). Given that T2D, itself, is likely to have been an advantageous phenotype only via pleiotropic variants acting through beneficial traits, we tested for association of index SNVs at distinct T2D signals with phenotypes available in the UK Biobank *(47)* (**Supplementary Materials and Methods, Figure S19**). We found that T2D risk alleles that were also associated with increased weight (and other obesity-related traits) tended to have more recent origin when compared to the genome-wide mutation age distribution at the same derived allele frequency (*p*<0.05 in all African ancestry populations), consistent with positive selection (**Figure S18**). Excluding these SNVs removed the selection signature that was observed in African ancestry populations. These observations are consistent with positive selection of T2D risk alleles that has been driven by the promotion of energy storage and usage appropriate to the local environment *(48)*. Further work is needed to characterise the specific pathways responsible for this adaptation.

## Discussion

The worldwide prevalence of diabetes mellitus has quadrupled over the last 30 years *(49)* and was estimated by the International Diabetes Federation to affect 415 million adults in 2015. China and India have emerged as major epicentres of the disease *(50)*, and in the USA, T2D and the downstream complications disproportionately affect individuals from ethnic minorities *(51,52)*. In consideration of the global burden of the disease, the DIAMANTE Consortium has assembled the most ancestrally diverse collection of T2D GWAS to date. To maximise the potential of these valuable resources to understand the genetic contribution to T2D across populations, we have also developed robust protocols for trans-ancestry analyses that will be relevant to any complex human trait. We previously implemented a powerful meta-regression approach *(13)* to enable aggregation of GWAS summary statistics across diverse populations that allows for heterogeneity in allelic effects on disease risk that is correlated with ancestry. Our study highlighted the advantages of applying this approach to ancestrally diverse GWAS in DIAMANTE for: (i) discovery of association signals that are shared across populations by reducing the genomic control inflation due to residual stratification; (ii) understanding the extent and source of heterogeneity in allelic effects at distinct association signals; (iii) allowing for LD-driven heterogeneity to improve fine-mapping resolution; and (iv) deriving population-specific weights to improve transferability of trans-ancestry GRS.

It is essential that the human genetics research community takes advantage of the opportunities offered by the increasing availability of GWAS across ancestry groups and continues to bolster collections in underrepresented populations. In contrast to GWAS, multi-omics resources that are critical for the inference of effector genes and causal mechanisms through which complex trait association signals are mediated remain deficient for the very populations with the greatest burden of disease. Further methodological development is also critical to: (i) allow for increased levels of admixture within populations; enable fine-mapping of multiple causal variants at the same locus without the need for conditional analyses; and (iii) conduct trans-ancestry polygenic risk scores that take account of genome-wide SNVs because of different LD structure across diverse populations. Prioritising research in diverse populations to address these challenges will ultimately provide a more comprehensive and refined view of the genetic contribution to complex human traits, improving understanding of the molecular and biological processes underlying common diseases, and offering the most promising opportunities for clinical translation of GWAS findings to improve global public health.

## Data Availability

Association summary statistics from the meta-analysis are not currently available.

## ACKNOWLEDGEMENTS

A full list of acknowledgments and funding appears in **Supplementary Note 2**.

## Author contributions

<u>DIAMANTE Consortium co-ordination:</u> A.Mahajan, M.I.M., A.P.M. <u>Manuscript preparation:</u> A.Mahajan, C.N.S., W.Zhang, M.C.Y.N., L.E.P., H.K., G.Z.Y., S.Rüeger, L.S., A.L.G., M.B., J.I.R., M.I.M., A.P.M. <u>Co-ordination of ancestry-specific GWAS collections:</u> A.Mahajan, C.N.S., W.Zhang, M.C.Y.N., L.E.P., D.W.B., J.E.B., J.C.C., X.S., M.B. <u>Central analysis group:</u> A.Mahajan, C.N.S., W.Zhang, M.C.Y.N., L.E.P., H.K., Y.J.K., M.Horikoshi, J.M.M., D.T., S.Moon, S.-H.K., N.R.R., N.W.R., M.Loh, B.-J.K., J.B.M., K.L.M., J.E.B., J.C.C., X.S., M.B., J.I.R., M.I.M., A.P.M. *<u>PROX1</u>* <u>functional analyses:</u> G.Z.Y., F.A., J.M.T., A.L.G. <u>GRS analyses in FinnGen:</u> S.Rüeger, P.d.B.P. <u>Selection analyses:</u> L.S., S.R.M. <u>Single cell chromatin accessibility data:</u> J.Chiou, D.G., S.P., M.Sander, K.J.G. <u>Islet promoter HiC data generation:</u> I.M.-E., J.F. <u>Study-level primary analyses:</u> A.Mahajan, C.N.S., W.Zhang, M.C.Y.N., L.E.P., Y.J.K., M.Horikoshi, J.M.M., D.T., S.Moon, S.-H.K., K.Lin, F.B., M.H.P., F.T., J.N., X.G., A.Lamri, M.N., R.A.S., J.-J.L., A.H.-G., M.Graff, J.-F.C., E.J.P., J.Y., L.F.B., Y.T., Y.H., V.S., J.P.C., M.K., N.G., E.M.S., I.P., T.S., M.W., C.Sarnowski, C.G., D.N., S.Trompet, J.Long, M.Sun, L.T., W.-M.C.,M.Ahmad, R.N., V.J.Y.L., C.H.T.T., Y.Y.J., C.-H.C., L.M.R., C.Lecoeur, N.M.M., B.P.P., A.N., L.R.Y., G.C., R.A.J., S.Tajuddin, E.K., P.A., A.H.X., H.S.C., B.E.C., J.Tan, X.S., A.P.M. <u>Study-level phenotyping, genotyping and additional analyses:</u> Y.Y.J., L.S.A., A.A., C.A.A.-S., M.Akiyama, S.S.A., A.B., Z.B., J.B.-J., I.B., J.A.B., C.M.B., T.B., M.Canouil, J.C.N.C, L.-C.C., M.L.C., J.Chen, S.-H.C., Y.-T.C., Z.C., C.C., L.-M.C., M.Cushman, S.K.D., H.J.d.S., G.D., L.D., A.P.D., S.D., Q.D., K.-U.E., L.S.E., D.S.E., M.K.E., K.F., J.S.F., I.F., M.F., O.H.F., T.M.F., B.I.F., C.F., P.G., H.C.G., V.G., C.G.-V., M.E.G.-V., M.O.G., P.G.-L., M.Gross, Y.G., S.Hackinger, S.Han, A.T.H., C.H., A.-G.H., W.Hsueh, M.Huang, W.Huang, Y.-J.H., M.Y.H., C.-M.H., S.I., M.A.I., M.Ingelsson, M.T.I., M.Isono, H.-M.J., F.J., G.J., J.B.J., M.E.J., T.J., Y.K., F.R.K., A.Kasturiratne, T.Katsuya, V.K., T.Kawaguchi, J.M.K., A.N.K., C.-C.K., M.G.K., K.K., J.Kriebel, F.K., J.Kuusisto, K.Läll, L.A.L., M.-S.L., N.R.L., A.Leong, L.Li, Y.Li, R.L.-G., S.Ligthart, C.M.L., A.Linneberg, C.-T.L., J.Liu, A.E.L., T.L., J.Luan, A.O.L., X.L., J.Lv, V.L., V.M., K.R.M., T.M., A.Metspalu, A.D.M., G.N.N., J.L.N., M.A.N., U.N., I.N., Y.O., L.O., S.R.P., M.A.Pereira, A.P., F.J.P., B.P., G.Prasad, L.J.R.-T., A.P.R, M.R., R.R., K.R., C.Sabanayagam, K.Sandow, N.S., S.S., C.Schurmann, M.Shahriar, J.S., D.M.S., D.Shriner, J.A.S., W.Y.S., A.S., A.M.S., K.Strauch, K.Suzuki, A.T., K.D.T., B.Thorand, G.T., U.T., B.Tomlinson, F.-J.T., J.Tuomilehto, T.T.-L., M.S.U., A.V.-S., R.M.v.D., J.B.v.K., R.V., M.V., N.W.-R., E.W., E.A.W., A.R.W., K.W.v.D., D.R.W., Y.-B.X., C.S.Y., K.Yamamoto, T.Y., L.Y., K.Yoon, C.Y., J.-M.Y., S.Y., L.Z., W.Zheng. <u>Study-level principal investigator:</u> L.J.R., M.Igase, E.Ipp, S.Redline, Y.S.C., L.Lind, M.A.Province, C.L.H., P.A.P., E.Ingelsson, A.B.Z., B.M.P., Y.-X.W., C.N.R., D.M.B., F.M., Y.Liu, E.Z., M.Y., S.S.R., C.K., J.S.P., J.C.E., Y.-D.I.C., P.F., J.G.W., W.H.H.S., S.L.R.K., J.-Y.W., M.G.H., R.C.W.M., T.-Y.W., L.G., D.O.M.-K., G.R.C., F.S.C., D.B., G.Pare, M.M.S., H.A., A.A.M., X.-O.S., K.-S.P., J.W.J., M.Cruz, R.M.-C., H.G., C.-Y.C., E.P.B., A.D., E.-S.T., J.D., N.K., M.Laakso, A.Köttgen, W.-P.K., C.N.A.P., S.Liu, G.A., J.S.K., R.J.F.L., K.E.N., C.A.H., J.C.F., D.Saleheen, T.H., O.P., R.M., C.Langenberg, N.J.W., S.Maeda, T.Kadowaki, J.Lee, I.Y.M., R.G.W., K.Stefansson, J.B.M., K.L.M., D.W.B., J.C.C., M.B., J.I.R., M.I.M., A.P.M.

## LIST OF SUPPLEMENTARY MATERIALS

Supplementary Materials and Methods

Supplementary Note 1: Summary of loci identified through recent ancestry-specific and trans-ancestry meta-analyses incorporating GWAS from the DIAMANTE Consortium

Supplementary Note 2: Acknowledgements and funding

Supplementary Note 3: Contributors to FinnGen Figures S1 – S20

Tables S1 – S23

